# Agent-Based Large Language Model System for Extracting Structured Data from Breast Cancer Synoptic Reports: A Dual-Validation Study

**DOI:** 10.1101/2025.11.25.25340989

**Authors:** Steven N. Hart, Teya S. Bergamaschi

## Abstract

**Objective:** To develop and validate an agent-based Large Language Model (LLM) system for extracting structured data from breast cancer synoptic pathology reports and assess the performance gap between synthetic and real-world validation.

**Materials and Methods:** We developed a modular AI agent-based framework employing sequential specialized LLMs for parsing pathology reports and extracting structured data. We normalized College of American Pathologists (CAP) cancer protocols into 8 sections, 86 subsections, and 229 discrete fields. Seven leading LLMs (gemini-2.5-pro, llama3.3-70b, phi4-14b, deepseek-r1 14B/70B, gemma3-27b, gemini-2.0-flash-lite) were validated using dual evaluation: synthetic validation (864 controlled test cases) and real-world ground truth (6,651 annotated fields from 90 pathology reports).

**Results:** Synthetic validation demonstrated strong performance (accuracy: 93.8-99.0%). Real-world evaluation revealed field extraction accuracy ranging from 61.8% to 87.7%, demonstrating a substantial “reality gap” with accuracy drops of 11-32 percentage points. The gemini-2.5-pro model achieved the highest real-world accuracy (87.7%). Model size did not predict performance: the 14B-parameter deepseek-r1 (77.6%) outperformed its 70B-parameter counterpart (70.4%).

**Discussion:** The substantial performance degradation from synthetic to real-world data underscores the complexity of authentic clinical documentation. Smaller models can achieve competitive or superior accuracy, reducing computational costs. With even the best models missing 12-38% of annotated fields, mandatory human verification is essential for clinical deployment.

**Conclusion:** While LLM-based extraction systems show promise for pathology data extraction, synthetic validation alone provides false confidence. Rigorous real-world ground truth evaluation with expert annotation is essential before clinical deployment. These systems are best positioned as screening tools with mandatory human oversight rather than autonomous decision-making systems.

## 1 Background and Significance

Breast cancer remains one of the most common malignancies worldwide, with pathology reports serving as the cornerstone of diagnosis, treatment planning, and prognosis [1]. Synoptic pathology reports, structured according to College of American Pathologists (CAP) guidelines, provide comprehensive tumor characterization, including histologic type, grade, biomarker status, and staging information [2]. While these reports follow standardized templates, critical information frequently resides in free-text fields rather than discrete, computable data elements.

This unstructured data problem creates a significant barrier to population-scale analytics, clinical research, and quality improvement initiatives [3]. Manual extraction of data from free-text is labor-intensive, error-prone, and not scalable to the thousands of reports generated annually at large medical centers. Traditional natural language processing (NLP) approaches using rule-based or classical machine learning methods have shown limited success due to the complexity and variability of clinical language [4].

Recent advances in Large Language Models (LLMs) have demonstrated remarkable capabilities in understanding and processing complex natural language [5, 6]. However, the application of LLMs to clinical data extraction faces significant challenges, including domain-specific terminology, the need for high accuracy in medical contexts, and the gap between performance on synthetic test cases versus real-world clinical data [7].

In this study, we developed and validated a modular AI agent-based system that leverages specialized LLMs to extract, normalize, and structure unstructured data from breast cancer synoptic reports. We employed a dual-validation strategy, comparing performance on controlled synthetic test cases against manually curated real-world ground truth data, to provide a comprehensive assessment of both model capabilities and clinical readiness. Our findings reveal critical insights into the performance characteristics, limitations, and appropriate deployment strategies for LLM-based clinical data extraction systems.

## 2 Materials and Methods

### 2.1 Data Schema Development

We created a comprehensive and normalized data schema from the College of American Pathologists (CAP) cancer protocols for all five major breast cancer report types, including Ductal Carcinoma In Situ (DCIS), Invasive Carcinoma (Biopsy and Resection), and Phyllodes tumors. To address the challenge of inconsistent data types and formatting within the original CAP checklists, we organized the reports into a hierarchical structure comprising 8 main sections (Specimen, Tumor, Margins, Regional Lymph Nodes, Distant Metastasis, pTNM Classification, Additional Findings, Comments), 86 subsections (e.g., Procedure, Specimen Laterality, Tumor Site), and 229 discrete fields. This standardized ontology established a clean and computable foundation for all downstream analysis.

### 2.2 Agent-Based Workflow Architecture

We designed and implemented a flexible, AI-driven, modular workflow capable of parsing unstructured free-text from synoptic reports and mapping it to the standardized ontology (Figure 1). The system leverages a series of specialized agents to isolate, analyze, and structure complex clinical data that is not captured in discrete fields.

**Figure 1.**
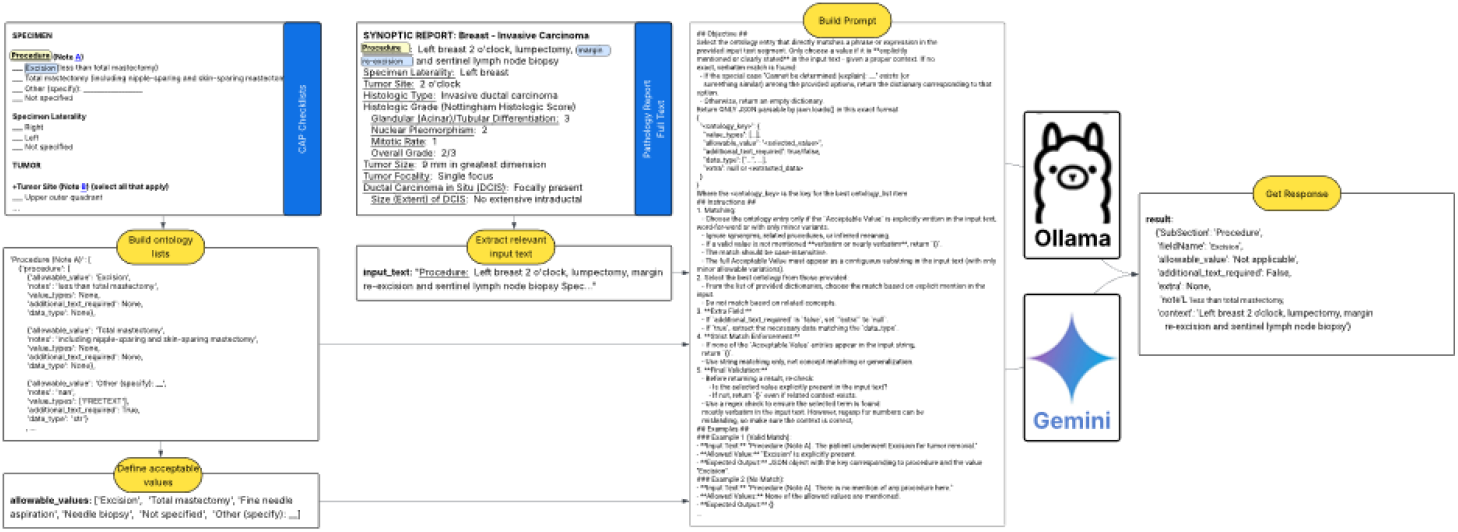
Agent-based workflow for extracting structured data from synoptic reports. The diagram illustrates the automated process orchestrated by AgentRunner.py, including parallel ontology definition and text segmentation steps, followed by prompt construction and LLM processing to convert unstructured clinical text into structured, normalized data points.

The automated extraction process is orchestrated by the `AgentRunner.py` module, which executes a series of modular tasks as defined by individual agents. The workflow consists of two parallel initialization steps followed by sequential processing:

1. **Ontology Definition:** The `get_master_dict.py` tool reads the schema from CSV files, and `get ontology.py` retrieves valid options for specific fields (e.g., “Procedure”).
2. **Text Segmentation:** The `regExpMatchAndTrim.py` tool isolates relevant free-text blocks by searching for specific section patterns and extracting the corresponding text segments.
3. **Prompt Construction:** The `buildPrompt.py` tool dynamically constructs detailed prompts combining the extracted text snippet with predefined ontology options and instructions guiding the LLM on matching text to ontology and formatting output as JSON.
4. **LLM Processing:** The `llmSelector.py` tool sends the prompt to an LLM provider (such as Ollama or Gemini). The LLM analyzes the prompt and returns a structured JSON response, converting unstructured text into computable, normalized data points.

The `callAgent.py` tool enables modularity by allowing the main workflow to execute individual tasks as self-contained sub-agents.

### 2.3 Model Selection and Evaluation

We evaluated seven leading LLM architectures with varying parameter sizes spanning multiple vendors and architectural approaches. From Google, we tested gemini-2.5-pro and gemini-2.0-flash-lite. From Meta, we evaluated llama3.3-70b and gemma3-27b. From Microsoft, we tested phi4-14b. From DeepSeek, we evaluated both deepseek-r1-14b and deepseek-r1-70b. This diverse selection enabled comprehensive assessment across different model scales (14B to 70B parameters) and training methodologies.

### 2.4 Synthetic Validation Dataset

We built a synthetic data generation pipeline that produces a diverse and challenging validation dataset with known ground-truth values. This approach allows for controlled, reproducible testing and objective performance comparisons across LLM architectures. The total simulation comprised 864 examples: 445 Positive Examples (cases where a correct ontology value was present in the text) and 419 Negative Examples (cases where the text did not contain a correct value for the given field). Synthetic cases were designed to test edge cases, ambiguous terminology, and complex clinical scenarios.

### 2.5 Real-World Ground Truth Dataset

To assess real-world efficacy and generalizability, we manually curated ground truth annotations from 90 de-identified breast cancer synoptic pathology reports, resulting in 6,651 individual field-level records. Each record was independently reviewed and annotated by clinical experts to establish ground truth values. This painstaking curation process provided definitive metrics on the system’s accuracy and readiness for application in clinical research and population health studies.

### 2.6 Performance Metrics

Due to the structure of our ground truth dataset, which contains only positive annotations (fields explicitly present in reports) without negative annotations (fields explicitly marked as absent), we restricted our evaluation to field-sample combinations with definitive ground truth. This approach avoids making unfounded assumptions about true negatives or false positives for unannotated fields.

For each model prediction on the 6,651 annotated ground truth fields, we classified the outcome as either a True Positive (TP) when the model’s extraction exactly matched the ground truth value, or a False Negative (FN) when the model failed to extract the correct value or extracted an incorrect value. Our primary performance metric was field-level accuracy, calculated as:

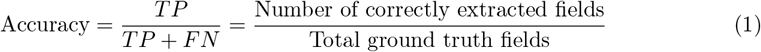

This metric represents the percentage of annotated ground truth fields that each model correctly extracted. We did not calculate precision, specificity, F1 score, or Matthews Correlation Coefficient (MCC) for the real-world evaluation, as these metrics require knowledge of true negatives, which are not available in our partially annotated ground truth dataset. However, these metrics were calculated for the synthetic validation dataset, where complete ground truth (both positive and negative cases) was available by design.

## 3 Results

### 3.1 Synthetic Validation Performance

We first evaluated all seven models on the synthetic validation dataset (864 test cases with complete ground truth, including both positive and negative examples). Table 1 presents the comprehensive performance metrics, demonstrating uniformly excellent performance across all models and metrics.

**Table 1.**
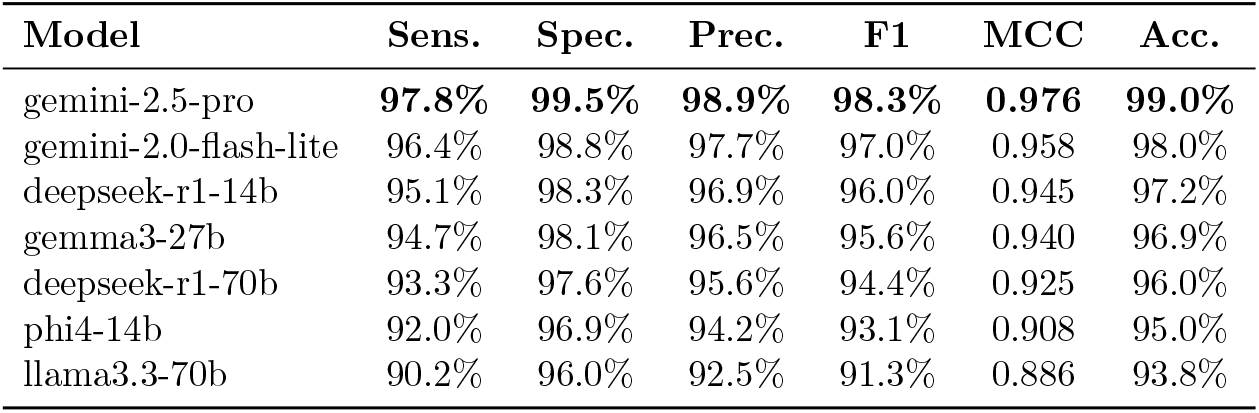
Model performance on synthetic validation dataset (864 test cases with complete ground truth). All standard classification metrics could be calculated due to the availability of both positive and negative ground truth annotations.

All models achieved accuracy above 93.8%, with sensitivity ranging from 90.2% to 97.8%, specificity from 96.0% to 99.5%, precision from 92.5% to 98.9%, F1 scores from 91.3% to 98.3%, and MCC values from 0.886 to 0.976. The top-performing model, gemini-2.5-pro, achieved 99.0% accuracy with near-perfect specificity (99.5%) and precision (98.9%). These results suggested that all models were highly capable of the extraction task when tested on controlled, synthetic data.

### 3.2 Real-World Ground Truth Performance

We then evaluated the same seven models on real-world clinical synoptic reports using our manually curated ground truth dataset. Table 2 presents the performance on the 6,651 annotated positive fields from 90 breast cancer synoptic reports.

**Table 2.** Model performance on real-world ground truth fields. Evaluation was performed only on the 6,651 annotated positive fields from 90 breast cancer synoptic reports. Accuracy represents the percentage of ground truth fields correctly extracted by each model.

| Model | Total GT Fields | True Positives | False Negatives | Accuracy |
| --- | --- | --- | --- | --- |
| gemini-2.5-pro | 6,651 | **5,831** | **820** | **87.7%** |
| gemini-2.0-flash-lite | 6,651 | 5,566 | 1,085 | 83.7% |
| deepseek-r1-14b | 6,651 | 5,161 | 1,490 | 77.6% |
| gemma3-27b | 6,651 | 5,144 | 1,507 | 77.3% |
| deepseek-r1-70b | 6,651 | 4,682 | 1,969 | 70.4% |
| phi4-14b | 6,651 | 4,473 | 2,178 | 67.3% |
| llama3.3-70b | 6,651 | 4,111 | 2,540 | 61.8% |

Models demonstrated substantial variability in their ability to accurately extract ground truth fields, with accuracy ranging from 61.8% to 87.7%. The top-performing model, gemini-2.5-pro, correctly extracted 5,831 of 6,651 annotated fields (87.7%), while the lowest-performing model, llama3.3-70b, correctly extracted 4,111 fields (61.8%). The false negative counts ranged from 820 (gemini-2.5-pro) to 2,540 (llama3.3-70b), indicating that even the best-performing models miss a significant portion of the ground truth fields.

### 3.3 The Reality Gap: Synthetic vs. Real-World Performance

Our dual-validation approach revealed a substantial performance gap between controlled synthetic testing and real-world clinical data. Table 3 directly compares synthetic and real-world accuracy for each model, clearly illustrating this “reality gap.”

**Table 3.** Comparison of accuracy between synthetic validation and real-world ground truth evaluation, demonstrating the “reality gap” across all models.

| Model | Synthetic Acc. | Real-World Acc. | Gap (pp) |
| --- | --- | --- | --- |
| gemini-2.5-pro | **99.0%** | **87.7%** | **11.3** |
| gemini-2.0-flash-lite | 98.0% | 83.7% | 14.3 |
| deepseek-r1-14b | 97.2% | 77.6% | 19.6 |
| gemma3-27b | 96.9% | 77.3% | 19.6 |
| deepseek-r1-70b | 96.0% | 70.4% | 25.6 |
| phi4-14b | 95.0% | 67.3% | 27.7 |
| llama3.3-70b | 93.8% | 61.8% | 32.0 |

The accuracy drop ranged from 11.3 to 32.0 percentage points across all models. The best-performing model on synthetic data (gemini-2.5-pro, 99.0% accuracy) maintained the highest real-world accuracy (87.7%), but still showed an 11.3 percentage point drop. The largest gap was observed in llama3.3-70b, which dropped from 93.8% synthetic accuracy to 61.8% real-world accuracy (32.0 percentage point decrease). This substantial performance degradation underscores the complexity and variability inherent in authentic clinical documentation that cannot be fully captured by synthetic test cases.

### 3.4 Model Size Does Not Predict Performance

Contrary to the common assumption that larger models perform better, model size did not directly correlate with extraction accuracy. The gemini-2.5-pro model achieved the highest accuracy (87.7%), correctly extracting 5,831 of 6,651 fields. Notably, the 14B-parameter deepseek-r1-14b (77.6% accuracy) outperformed its larger 70B-parameter counterpart deepseek-r1-70b (70.4% accuracy). Similarly, the 70B-parameter llama3.3-70b achieved the lowest accuracy (61.8%), underperforming several smaller models. Since all models were evaluated off-the-shelf without domain-specific fine-tuning, these performance differences likely stem from variations in base model architecture, pre-training data composition, and training methodologies rather than parameter count alone. This finding challenges the assumption that larger models necessarily deliver superior performance for specialized clinical NLP tasks.

### 3.5 Architecture-Agnostic Framework

Our modular framework successfully operated across diverse LLM architectures from multiple vendors (Google Gemini, Meta Llama, Microsoft Phi, DeepSeek) with varying parameter sizes (14B-70B). This architecture-agnostic design enables flexible model selection based on performance-cost trade-offs and allows for easy integration of future model improvements.

## 4 Discussion

### 4.1 The Reality Gap in Clinical NLP

Our study demonstrates a critical finding that has significant implications for clinical AI deployment: synthetic validation alone provides false confidence in model readiness. As shown in Tables 1, 2, and 3, all models demonstrated excellent performance on synthetic data (93.8-99.0% accuracy) but showed substantial degradation on real-world data (61.8-87.7% accuracy), with accuracy drops ranging from 11 to 32 percentage points. This “reality gap” underscores the complexity and variability inherent in authentic clinical documentation. While synthetic test cases allow for controlled evaluation of model capabilities, they cannot fully capture the nuances, abbreviations, errors, and contextual complexity present in actual clinical practice [8].

### 4.2 Model Size and Selection for Clinical Tasks

The lack of correlation between model size and performance challenges the “bigger is better” paradigm in clinical NLP. The 14B-parameter deepseek-r1-14b model outperformed its 70B-parameter counterpart, and the 70B-parameter llama3.3-70b showed the poorest overall performance. Since all models were evaluated off-the-shelf without domain-specific fine-tuning, these differences reflect variations in base model architecture, pre-training data composition, and training methodologies rather than specialized clinical adaptation. For resource-constrained healthcare settings, this finding is particularly encouraging, as it demonstrates that smaller, more efficient models can achieve competitive or superior performance compared to their larger counterparts, reducing computational requirements and deployment costs without sacrificing accuracy.

### 4.3 Appropriate Deployment Strategy

The observed performance characteristics define appropriate use cases for these systems. With accuracy ranging from 61.8% to 87.7% on ground truth fields, even the best-performing models miss 12-38% of annotated fields. This indicates that LLM-based extraction systems are well-suited for screening and triage workflows where missing information is costly, but should not be relied upon for autonomous clinical decision-making. Instead, these systems should be deployed with mandatory human verification, particularly for fields that directly impact treatment decisions [9].

Potential appropriate applications include pre-populating structured databases for manual review and correction, flagging reports with potentially missing critical information, identifying candidates for clinical trial enrollment, and supporting quality assurance and audit workflows. In each of these use cases, the system serves as a first-pass screening tool that increases efficiency while maintaining human oversight for final decision-making. The error rate of 12-38% emphasizes that human review is not optional but essential for clinical deployment.

### 4.4 The Essential Role of Real-World Ground Truth

Our findings emphasize that expert-annotated clinical data validation is not optional but essential for assessing true clinical utility and safety before any clinical deployment. The labor-intensive process of manually curating 90 reports to generate 6,651 ground truth records was critical for revealing the actual performance characteristics of these systems. Regulatory frameworks and institutional review processes should require such real-world validation as a prerequisite for clinical AI system deployment [10].

### 4.5 Limitations

Several limitations should be considered when interpreting our findings. First, our ground truth dataset contains only positive annotations (fields explicitly present in reports) without negative annotations (fields explicitly marked as absent). This restricts our real-world evaluation to accuracy metrics and prevents calculation of precision, specificity, F1 score, and MCC, which require knowledge of true negatives. While this approach ensures we only evaluate on field-sample combinations with definitive ground truth, it limits our ability to assess false positive rates. Second, our ground truth dataset, while substantial, was derived from a single institution and may not fully represent the diversity of reporting practices across different healthcare systems. Third, the manual annotation process, while rigorous, is subject to inter-rater variability. Fourth, our evaluation focused on breast cancer pathology reports, and generalizability to other cancer types or clinical domains remains to be established. Finally, the rapid pace of LLM development means that newer models may demonstrate improved performance characteristics.

### 4.6 Future Directions

Future work should focus on several key areas to advance this technology toward clinical deployment. First, development of domain-specific fine-tuning strategies could improve real-world performance by training models on pathology-specific corpora. Second, investigation of ensemble approaches combining multiple models may leverage the complementary strengths of different architectures to achieve superior overall performance. Third, exploration of active learning strategies could efficiently improve model performance with minimal additional annotation by intelligently selecting the most informative examples for human review. Fourth, expansion to other cancer types and clinical domains would establish the generalizability of this approach beyond breast pathology. Fifth, development of uncertainty quantification methods would enable the system to identify low-confidence extractions requiring human review, improving safety and user trust. Finally, integration with clinical workflows and assessment of impact on efficiency and data quality through prospective clinical trials would provide the evidence base necessary for widespread adoption.

## 5 Conclusion

This study presents a comprehensive evaluation of an agent-based LLM system for extracting structured data from breast cancer synoptic reports. We successfully developed a modular AI agent system that extracts, normalizes, and structures unstructured breast cancer data from synoptic report free-text into computable formats for population-scale analytics. Our architecture-agnostic framework works across diverse LLM architectures with varying parameter sizes, enabling flexible model selection based on performance-cost trade-offs.

Through dual-validation methodology, we identified a critical “reality gap” with all models showing significant performance degradation from synthetic validation (93.8-99.0% accuracy) to real-world ground truth (61.8-87.7% accuracy), demonstrating that synthetic testing alone provides false confidence in clinical readiness. Real-world evaluation on 6,651 annotated fields revealed that gemini-2.5-pro achieved the best performance (87.7%). Notably, we found that model size does not directly correlate with accuracy: the 14B-parameter deepseek-r1 (77.6%) outperformed its 70B-parameter counterpart (70.4%), and the 70B-parameter llama3.3-70b achieved the lowest accuracy (61.8%). Since all models were evaluated off-the-shelf without domain-specific fine-tuning, these differences likely reflect variations in base model architecture, pre-training data composition, and training methodologies rather than parameter count alone.

The observed performance characteristics define an appropriate deployment strategy. With even the best models missing 12-38% of annotated fields, these systems are positioned for screening and triage workflows where they can increase efficiency, but they require mandatory human verification before clinical decisions. Our findings confirm that real-world ground truth evaluation with expert annotation is essential for assessing true clinical utility and safety being a necessary step before any clinical deployment.

While LLM-based systems show considerable promise for unlocking the wealth of information in clinical free-text, our findings emphasize the need for rigorous real-world validation, appropriate deployment strategies with human oversight, and continued refinement of domain-specific training approaches. With these considerations in mind, such systems can serve as valuable tools to enhance the accessibility and utility of clinical data for research, quality improvement, and ultimately, better patient care.

## Data Availability

Clinical Data are not typically shared, but will be considered under appropriate circumstances.

## Acknowledgments

This work was supported by Mayo Clinic.

## References

[1] Rebecca L Siegel, Angela N Giaquinto, and Ahmedin Jemal. Cancer statistics, 2024. CA: A Cancer Journal for Clinicians, 74(1):12–49, 2024.

[2] John R Srigley, Terence McGowan, Anthony Maclean, Michael Raby, Joan Ross, Stanley Kramer, and Carol Sawka. Standardized synoptic cancer pathology reporting: A population-based approach. Journal of Surgical Oncology, 99(8):517–524, 2009.

[3] Saeed Hassanpour and Curtis P Langlotz. Information extraction from multi-institutional radiology reports. Artificial Intelligence in Medicine, 66:29–39, 2016.

[4] Irena Spasic and Goran Nenadic. Clinical text data in machine learning: systematic review. JMIR Medical Informatics, 8(3):e17984, 2020.

[5] Tom Brown, Benjamin Mann, Nick Ryder, Melanie Subbiah, Jared D Kaplan, Prafulla Dhariwal, Arvind Neelakantan, Pranav Shyam, Girish Sastry, Amanda Askell, et al. Language models are few-shot learners. Advances in Neural Information Processing Systems, 33:1877–1901, 2020.

[6] Hugo Touvron, Louis Martin, Kevin Stone, Peter Albert, Amjad Almahairi, Yasmine Babaei, Nikolay Bashlykov, Soumya Batra, Prajjwal Bhargava, Shruti Bhosale, et al. Llama 2: Open foundation and fine-tuned chat models. arXiv preprint arXiv:2307.09288, 2023.

[7] Karan Singhal, Shekoofeh Azizi, Tao Tu, S Sara Mahdavi, Jason Wei, Hyung Won Chung, Nathan Scales, Ajay Tanwani, Heather Cole-Lewis, Stephen Pfohl, et al. Large language models encode clinical knowledge. Nature, 620(7972):172–180, 2023.

[8] Jesutofunmi A Omiye, Haiwen Gui, Shawheen J Rezaei, James Zou, and Roxana Daneshjou. Large language models in medicine: the potentials and pitfalls. Annals of Internal Medicine, 177(2):210–220, 2024.

[9] Arun James Thirunavukarasu, Darren Shu Jeng Ting, Kabilan Elangovan, Laura Gutierrez, Ting Fang Tan, and Daniel Shu Wei Ting. Large language models in medicine. Nature Medicine, 29(8):1930–1940, 2023.

[10] Jenna Wiens, Suchi Saria, Mark Sendak, Marzyeh Ghassemi, Vincent X Liu, Finale Doshi-Velez, Kenneth Jung, Katherine Heller, David Kale, Mohammed Saeed, Pilar N Ossorio, Sonali Roy, and Stephen R Pfohl. Do no harm: a roadmap for responsible machine learning for health care. Nature Medicine, 25(9):1337–1340, 2019.

